# Molecular characterisation of a *Klebsiella pneumoniae* neonatal sepsis outbreak in a rural Gambian hospital: a retrospective genomic epidemiology investigation

**DOI:** 10.64898/2026.03.03.26347025

**Authors:** Ebenezer Foster-Nyarko, Alasana Bah, Williams Oluwatosin Adefila, Isaac Osei, Ousman Barjo, Rasheed Salaudeen, Shola Able-Thomas, Mariama Jammeh, Abdoullah Nyassi, Erkison Ewomazino Odih, Kathryn Elizabeth Holt, Abdul Karim Sesay, Grant A. Mackenzie

## Abstract

**Background:** *Klebsiella pneumoniae* is a common cause of neonatal sepsis in Africa, and is frequently hospital acquired. We recently reported an outbreak of multidrug-resistant *K. pneumoniae* sepsis amongst neonates at a rural hospital in The Gambia, West Africa, involving 57 cases and case fatality of 60%. Here we undertook a retrospective pathogen genomic epidemiology study of clinical and environmental *K. pneumoniae* isolated during the outbreak, to identify the outbreak strain, refine the epidemic curve, confirm the environmental sources of contamination, monitor control of the outbreak, and characterise the outbreak strain in the context of the local and global pathogen population.

**Methods and Findings:** We sequenced all blood culture isolates identified as *K. pneumoniae* from patients aged 0–59 months (n=51 available, 77% from neonates), together with *K. pneumoniae* cultured from environmental samples during the outbreak investigation (n=16), and 56 stored blood culture surveillance isolates available from the previous decade (34 from neonates). Sequencing was performed using Oxford Nanopore Technologies (ONT) Mk10 flowcells and a PromethION instrument, yielding mostly complete genomes (79%). Genomic analysis revealed 23% of isolates were *K. quasipneumoniae* and identified the outbreak strain as *K. pneumoniae* ST39 with capsular (K) locus KL62. This strain was responsible for 29 cases (16 fatalities) and was identified in three samples of intravenous fluids collected from the neonatal ward during the investigation. It harboured a ∼187 kbp IncF plasmid carrying the extended-spectrum beta-lactamase (ESBL) gene *bla*_CTX-M-15_ and *aac*(3)-*IId*, encoding resistance to third-generation cephalosporins and gentamicin, respectively. The outbreak strain was not identified amongst historical surveillance isolates, and it was distinct from a KL23-ST39 strain responsible for an earlier outbreak at the Sir Edward Francis Small Teaching Hospital in Banjul, the country’s capital 7 years prior. Comparing the outbreak strain with publicly available genome data for ST39 and its associated sublineage (SL) 39, we observed SL39 has diversified into three common clonal groups, each associated with multiple K types, that have spread across Africa, Asia and Europe and have been associated with outbreaks of neonatal sepsis in Africa and elsewhere. We find SL39 is typically multidrug resistant, however the specific ESBL and carbapenemase genes vary by geographic location.

**Conclusions:** Pathogen whole-genome sequencing refined our understanding of the outbreak, allowing more precise identification to refine case numbers and case fatality calculations, and for precise identification of multi-use intravenous fluid bags as the source of the outbreak despite other samples being culture-positive for unrelated *K. pneumoniae*. This highlights the importance of infection prevention and control in reducing neonatal fatalities in low-resource settings, and the critical risk associated with multi-use reagents and equipment when caring for vulnerable neonates. The genomic analysis enabled us to identify and characterise the outbreak strain at high resolution, and together with global data highlights SL39 as an emerging high-risk multidrug-resistant, globally distributed clone of *K. pneumoniae,* capable of sustained transmission and high fatality.

**Impact Statement:** Neonatal sepsis caused by *Klebsiella pneumoniae* kills an estimated 800,000 infants annually, with the greatest burden in sub-Saharan Africa where multidrug-resistant strains are increasingly prevalent. Yet genomic investigations of outbreak strains from rural African settings remain scarce, limiting our understanding of transmission dynamics and the global emergence of high-risk lineages. In this study, we apply whole-genome sequencing to a multidrug-resistant *K. pneumoniae* ST39 neonatal sepsis outbreak at a rural Gambian hospital, demonstrating how pathogen genomics can decisively refine outbreak investigations in low-resource settings: it corrected case counts, pinpointed contaminated multi-use intravenous fluid bags as the primary source, and revealed a novel ISKpn14-mediated chromosomal resistance island. Contextualising the outbreak strain within a global collection of 684 ST39 genomes, we identify sublineage 39 as a diversified, globally distributed, multidrug-resistant clone whose three dominant clonal groups have caused neonatal sepsis outbreaks across Africa, Asia, and Europe. Our findings reinforce the urgent need for sustained investment in genomic surveillance and infection prevention infrastructure in African neonatal units, and provide actionable data to guide targeted screening and vaccine design against prevalent African *K. pneumoniae* lineages.

**Data summary:** The raw sequencing data generated for this study have been deposited in the NCBI Sequence Read Archive (SRA) under BioProject accession PRJNA1367967. Individual isolate sequencing data are provided in **Supplementary File 3** and are available under BioSample accessions SAMN53359187 through SAMN53359276, with corresponding SRA accessions SRR36200451 through SRR36200540.

The complete genome assembly for a representative ST39 outbreak isolate, 38833B1 is available in GenBank under BioSample accession SAMN53359242, including the complete sequence of pNS39_A under GenBank accession PX620516.

Code used to generate the figures and reproduce the analyses presented in this study is available at https://github.com/efosternyarko/molecular-characterisation-klebsiella-pneumoniae-neonatal-sepsis-outbreak-gambia.

## Introduction

*Klebsiella pneumoniae* has emerged as the leading cause of neonatal sepsis globally, responsible for an estimated 800,000 annual neonatal deaths worldwide (1). The burden is disproportionately concentrated in sub-Saharan Africa, where neonatal mortality reaches 27 deaths per 1,000 live births, with infections accounting for a substantial proportion of deaths (2).

Treatment of *K. pneumoniae* sepsis is complicated by escalating antimicrobial resistance (AMR), and *K. pneumoniae* is estimated as one of the largest contributors to AMR-associated deaths globally across all age groups, and the largest contributor to neonatal deaths from AMR infections (3). The WHO-recommended first- and second-line neonatal sepsis therapies (ampicillin or third-generation cephalosporins (3GCs) such as cefotaxime or ceftriaxone, plus gentamicin) are now largely ineffective (4–6), contributing to case fatality of up to 77% in documented outbreaks (7–9).

*K. pneumoniae* are intrinsically resistant to ampicillin and frequently acquire resistance to 3GCs through acquired extended-spectrum β-lactamase (ESBL) genes. The latest WHO Global Antimicrobial Resistance and Use Surveillance System (GLASS) report (10) indicates over 55% of *K. pneumoniae* clinical isolates globally are now resistant to 3GCs, and the rate exceeds 70% in Africa. Studies in African neonatal care units indicate ∼80% of *K. pneumoniae* isolates are 3GC resistant, and >20% are also resistant to carbapenems (4, 5, 11).

Globally, ESBL- and carbapenemase-producing *K. pneumoniae* are associated with a small number of high-risk clones, as defined by molecular analysis (12–14) and linked to outbreaks across multiple continents. Pathogen genomic data from African neonates is relatively limited, but reports from Malawi and Kenya have identified diverse *K. pneumoniae* clones including some that are also recognised as high-risk to health in Europe and other continents (e.g., ST14, ST15, ST17, ST101, ST307, ST340) (15–19).

Sources of bacterial infection in neonatal care units have been conceptualised as either vertical transmission (acquired from colonised mothers) or horizontal transmission (acquired from the hospital environment, including contaminated surfaces, equipment, reagents and healthcare workers) (20–22). Outbreak investigation reports from low-resource African settings are sparse, but implicate horizontal transmission, particularly via contamination of multi-use equipment and reagents (23–25). Improving understanding of the sources of *K. pneumoniae* outbreaks in neonatal units in Africa could help inform improvements in infection prevention and control (IPC). Characterisation of the associated outbreak strains could help to identify problem clones with increased risk of transmission and/or mortality, to inform more targeted screening in African settings (e.g., screening for colonisation of neonates, hospital environment, or healthcare workers).

We recently reported an outbreak of multidrug-resistant *K. pneumoniae* sepsis amongst children under 5 years old at a rural hospital in The Gambia, West Africa (26). A total of 76 cases were identified, including 57 neonates, with overall case fatality (CF) of 56% (60% CF amongst neonates) (26). Environmental screening in the neonatal and labour wards yielded *K. pneumoniae* positive cultures from water, the ward environment, and intravenous fluid. Multiple measures were taken to improve IPC, which ultimately led to a decline in cases and culture-negative environmental screening, after which the outbreak was declared over.

Here, we report a retrospective genomic investigation of clinical and environmental *K. pneumoniae* isolated during the outbreak, aiming to (1) identify the outbreak strain and refine the epidemic curve, (2) confirm the primary environmental sources of contamination, and (3) characterise the outbreak strain in the context of the local and global *K. pneumoniae* population.

## Methods

### Study design and setting

Bansang Hospital is a secondary health facility in Central River Region of The Gambia with a 75-bed paediatric ward that includes a 30-bed neonatal ward (**Figure 1B**). The hospital serves as the primary healthcare facility for a catchment population of approximately 200,000. The outbreak occurred during a period of continual disease surveillance for pneumonia, sepsis, and meningitis in children aged 0-59 months as part of the Pneumococcal Vaccine Schedules trial (ISRCTN17145509) (27, 28). The genomic investigation focuses on the period from January 2023 to March, 2024, and includes all blood culture isolates identified as *K. pneumoniae* from this period. The authors of this study did not have access to any information that could potentially identify individual participants. Environmental isolates identified during the outbreak investigation, and historical *K. pneumoniae* clinical isolates from surveillance at Basse and Bansang hospitals from 2012-2022, were also sequenced to provide context for strain circulation patterns. All study specimens were processed at the MRC Unit The Gambia’s microbiology laboratory in Basse (**Figure 1B**), which maintains Good Clinical Laboratory Practice accreditation and participates in UK National External Quality Assessment Service proficiency testing programs.

### Case definitions

The original outbreak report details the case definitions and patient characteristics (26). Briefly, a case was defined as a patient aged 0 to 59 months admitted to Bansang Hospital from whom *K. pneumoniae* was isolated from blood between June 2023 and March 2024. Neonatal cases were defined as those who were aged 0-28 days at the time the blood was collected for culture. Case fatality was calculated as the proportion of confirmed cases who died before being discharged from the hospital.

### Microbiological methods

Clinical blood specimens were processed using standardised protocols and incubated in the BACTEC FX automated culture system (BD Diagnostics) at 37°C for five days. Upon detection of growth (indicated by system flagging and visible turbidity), positive blood culture bottles were subcultured onto MacConkey agar and Columbia agar supplemented with sheep blood, then incubated at 37°C in 5% CO_2_.

Following incubation, plates were examined for colony morphology characteristic of *Klebsiella* species. On blood agar, presumptive *Klebsiella* colonies appeared as shiny, mucoid, round colonies with a greyish-white appearance and diameter >3 mm. Strains displaying distinctive “cotton candy” or “stringy” morphology due to extensive capsular polysaccharide production were also selected. On MacConkey agar, lactose-fermenting colonies with smooth, mucoid, pink to reddish coloration were identified as potential *Klebsiella* isolates. Putative *Klebsiella* isolates were further characterised biochemically using API 20E, with results interpreted using the APIWEB website (https://apiweb.biomerieux.com).

### Environmental sampling strategy

Environmental sampling was conducted during October, November, and December 2023 using a systematic approach based on US CDC environmental sampling guidelines (29). Sampling sites were selected based on epidemiological investigation findings and included high-touch surfaces, patient care equipment, water sources, and intravenous fluid preparation areas. Full details of the samples taken, and the culture results, are given in the outbreak report (26).

Surface sampling employed sterile pre-moistened swabs for non-absorbent surfaces and composite sampling methods for large areas, as recommended for optimal pathogen recovery (30). Water samples were collected in sterile containers with sodium thiosulfate for chlorine neutralisation and transported at 4°C for processing within 24 hours. Intravenous fluid samples were collected aseptically from administration sets, infusion bags, and preparation areas. All environmental samples were processed using enrichment culture in peptone water followed by plating on MacConkey agar and Columbia agar supplemented with 5% sheep blood for *K. pneumoniae* detection. Species identification was performed using the Analytical Profile Index (API) biochemical identification system, as mentioned earlier. Environmental isolates underwent identical characterisation procedures as clinical isolates.

### Antimicrobial susceptibility testing

Antimicrobial susceptibility testing was performed using the Kirby-Bauer disc diffusion method, following Clinical and Laboratory Standards Institute (CLSI) guidelines (31). Isolates were tested against ampicillin, ceftazidime, cefotaxime, cefoxitin, ceftriaxone, chloramphenicol, ciprofloxacin, gentamicin, trimethoprim/sulfamethoxazole and tetracycline discs. Quality control was maintained using American Type Culture Collection (ATCC) reference strains including *K. pneumoniae* ATCC 700603 (ESBL-positive), *Escherichia coli* ATCC 25922 (susceptible control), and *Pseudomonas aeruginosa* ATCC 27853. All testing was performed by trained laboratory technologists with results reviewed by clinical microbiologists.

### Whole-genome sequencing and assembly

All isolates were sequenced using Oxford Nanopore Technologies (ONT) long-read sequencing at the Medical Research Council Unit the Gambia’s Core Genomics Facility in Fajara, The Gambia. DNA extraction was performed using the Qiagen genomic-tip 20/G DNA Kit (Qiagen) from overnight cultures. Library preparation was performed using the ONT rapid barcoding kit, following manufacturer’s guidelines. Libraries were loaded onto PromethION R10 flow cells and sequenced on the PromethION 2 Solo for 72 hours. Reads were basecalled using Dorado v0.7.2 with SUPer-accuracy model (SUP) v5.0, via the MRCG at LSHTM’s high-performance computing clusters. Average yield per isolate was 433,217 reads (range 6,540–2,106,200) and 1,346 bp (range 253–3,809). Genomes were assembled using Hybracter v0.7.3 (32) via the Cloud Infrastructure for Microbial Bioinformatics (CLIMB) platform (33), and assembly quality assessed using QUAST v5.0.0 [de6973bb] (34). All isolates yielded an assembly of the expected size (range, 5.13–5.95 Mbp). Most isolates (n=98, 78%) yielded a >5 Mbp contiguous chromosome sequence, with up to 11 total contigs (median 4) (full assembly statistics in **Table S2**).

A random selection of isolates belonging to the ST39 (n=13) and ST1788 (n=4) clusters were additionally sequenced using Illumina short-read sequencing. An aliquot of the same DNA extract used for ONT sequencing was used as input to Illumina library preparation, utilising the Nextera Flex DNA Library Prep Kit (Illumina). Libraries were subjected to 2×150 bp paired-end sequencing on Illumina NextSeq 500 platform. Raw sequencing data underwent quality assessment using FastQC (35) with adapter trimming performed using Trimmomatic (36), followed by read processing and error correction using FLASH v1.2.11 (https://github.com/ebiggers/flash) (37) to merge overlapping paired-end reads prior to assembly. Merged reads and remaining unmerged paired reads were both retained for subsequent assembly using SPAdes v3.15.5 (27) on CLIMB (33).

### Genomic typing and characterisation

Genome assemblies were uploaded to the online Pathogenwatch platform v2.3.1 (28), which assigns species based on mash distances to a curated set of reference genomes; utilises Kleborate v3.2.4 (38) for comprehensive genomic characterisation including multi-locus sequence typing (MLST) (39, 40), AMR variant calling, and virulence locus detection; Kaptive v3.0.0b6 (41, 42) for capsular (K) and lipopolysaccharide (O) locus typing; and assigns core-genome multi-locus sequence types (cgSTs) and LIN codes defined by the BIGSdb-Pasteur scheme (41, 43, 44). For *K. pneumoniae* genomes, we used Pathogenwatch to generate a pairwise genetic distance matrix which it calculates from a concatenated alignment of 1,972 core genes (2,172,367 bp). For comparison with public genome data, we used the Pathogenwatch platform to search for all genomes assigned to ST39, and downloaded the available meta-data (year, country) and genotyping results from Pathogenwatch to compare with our study data (i.e., all data were genotyped consistently using the same methods implemented in Pathogenwatch).

### Transmission cluster analysis

Putative transmission clusters were identified using single-linkage clustering of neonatal cases, implemented in the Transmission Estimator tool (45). Transmission clusters were defined as groups of isolates that satisfied two criteria: (i) Pathogenwatch genetic distance ≤10 from at least one other cluster member (this threshold corresponds to genome-wide SNP distance of 20-25, widely accepted for identifying nosocomial transmission clusters of *K. pneumoniae* (46)); and (ii) temporal distance, isolated within four weeks (28 days) of at least one other cluster member. The reliability of ONT-sequenced genomes for cluster analysis was assessed by comparing the distance matrices generated using ONT-based assemblies vs those generated using Illumina-based assemblies, for the 17 samples randomly selected from the ST39 (n=13) and ST1788 (n=4) clusters (**Supplementary File 2** and **Appendix A**).

### Plasmid assembly and comparative genomics

Complete plasmid sequences were assembled from long-read Oxford Nanopore sequencing data using Hybracter v0.7.3 (32). Plasmid replicon types were identified using Abricate v1.0.1 (47) with the PlasmidFinder database (updated 14 January 2025), and antimicrobial resistance genes were detected using the ResFinder database (updated 14 January 2025).

To identify closely related reference sequences, the outbreak-associated plasmid pNS39_A (derived from ST39 isolate 38833B1) was queried against the PLSDB (plasmid) database (48) (accessed 14 November 2025) using BLASTn. The top-scoring hit was selected as the reference sequence for detailed comparative analysis. Pairwise alignment between pNS39_A and the reference plasmid was performed using BLASTn with default parameters.

Alignment coverage was quantified using two complementary approaches: (i) the longest single contiguous alignment block, and (ii) cumulative coverage from multiple alignment blocks ranked by length. Coverage estimates, alignment block size distributions, sequence identity metrics, and cumulative coverage curves were visualised using custom Python v3.13.9 scripts with matplotlib 3.10.7 and pandas v2.2.3 libraries.

To assess plasmid conservation across the outbreak population, individual plasmid sequences from all ST39 isolates were aligned to reference plasmids pNS39_A (187,092 bp) and pNS39_B (66,878 bp) derived from representative outbreak strain, 38833B1, using minimap2 v2.28 (49) with the asm5 preset optimised for assembly-to-assembly comparisons. Alignments were converted to sorted BAM format using samtools v1.21 (50). Coverage statistics were calculated using samtools coverage, generating metrics including reference coverage (percentage of reference plasmid covered by query alignment), query coverage (percentage of query plasmid aligning to reference), mean and median sequencing depth, and sequence identity (calculated from edit distance using the NM:i tag in SAM format, representing mismatches, insertions, and deletions). Coverage heatmaps were generated using Python v3.13.9 with matplotlib v3.10.7, seaborn v0.13.2, and pandas v2.2.3 libraries.

### Chromosomal resistance gene integration analysis

Antimicrobial resistance genes were identified across all 32 assemblies using ABRicate v1.0.1 (47) with the ResFinder and CARD databases (≥80% nucleotide identity, ≥80% coverage). Insertion sequence (IS) elements were predicted using ISEScan v1.7.2.3 (51, 52) against the ISfinder reference database.

For each assembly, distances between resistance genes and IS elements on the same contig were calculated using a custom Python (v3.10) pipeline; IS elements within 10 kb of a resistance gene were classified as proximal, with position recorded as upstream, downstream, or overlapping. In isolate 38277B1, *bla*_CTX-M-15_ was identified on the chromosome rather than on a plasmid; flanking IS elements were extracted using BioPython v1.81 and submitted to the ISfinder BLASTn service (52) for definitive classification against 6,013 curated IS sequences. Target site duplications (TSDs) flanking each IS element were identified by manual inspection of the 20 bp immediately adjacent to both termini in the assembled sequence.

To determine whether the chromosomal integration replaced a pre-existing mobile element, the corresponding locus from reference isolate 38833B1 was extracted and gene cluster comparison was performed using clinker v0.0.32 (53) with a minimum protein identity threshold of 0.6; open reading frames in extracted loci were predicted with Pyrodigal v3.7.0 (54, 55). Genome annotation for regional context was performed with Bakta v1.8.1 (56) using database v5.1.

### Statistical and bioinformatics methods

All other data analysis and visualisation were conducted using R v4.3.0, using tidyverse v0.4.6 for data processing and ggplot2 v3.5.2 for visualisation. Phylogenetic trees were visualised using ggtree v3.16.3.

### Ethical considerations

This study was conducted under the ethical approval for the Pneumococcal Vaccine Surveillance trial obtained from the Gambia Government/MRC Joint Ethics Committee (reference SCC1577.v2, approved 22 February 2018) and the London School of Hygiene & Tropical Medicine Ethics Committee (reference #14515, approved 13 November 2019). The historical isolates were generated as part of an earlier surveillance study that was approved by the Gambia Government/MRC Joint Ethics Committee (ref SCC 1087, 17/8/2007). For both studies, written informed consent was obtained from each participant’s parent or guardian, and covered storage of samples for future tests and research relating to infection.

## Results

### Outbreak investigation and response

Details of the outbreak investigation and response have been published previously (26), key details are reproduced here to provide context to the genomic investigation. **Figure 1A** shows the previously reported epidemic curve from January 2023 to March 2024, with additional annotation of fatalities, breakdown into cases vs deaths, and showing the timing and results of environmental sampling. Between January 2023 and March 2024, 54 laboratory-cases were identified at Bansang Hospital, including 46 (85%) in neonates. Monthly case counts (all ages, i.e. 0-59 months) rose from 1–4 cases in January through May to 10–11 cases in June and July. Case fatality increased during the outbreak, rising from 0% (n=0/13) in January–May, to 54% (n=29/54) in June–December and 63% (n=29/46) in neonates. Most affected neonates were born in Bansang Hospital (n=34/46, 74%).

Initial investigation of the outbreak, including assessment of clinical and microbiological data and IPC risks, identified multiple IPC issues across both labour and neonatal wards and control measures were implemented in September (26). Systematic environmental sampling conducted in October identified contamination with *K. pneumoniae* throughout the neonatal unit and labour ward. Of 48 environmental samples collected, 14 (29%) yielded *K. pneumoniae* growth, while five samples grew coagulase-negative *Staphylococcus* (considered environmental contaminants) and one sample yielded *Pseudomonas* species (**Supplementary File 1**). *K. pneumoniae* contamination was detected in running water from taps in both the labour ward and neonatal ward, multiple delivery beds in the first delivery room (2/2 beds positive), and three (3/3) neonatal resuscitation beds. Additional contaminated sites included the weighing scale in the labour ward and station surfaces where drugs and medical tools were stored (**Supplementary Figure 1**).

Most significantly, all intravenous fluids used for medication dilution and maintenance tested positive for *K. pneumoniae* (4/4 samples positive). Phenotypic antimicrobial resistance profiling revealed the prevalence of multidrug resistance in both clinical and environmental isolates, with 10 of 11 clinical isolates (91%) and 7 of 14 environmental isolates (50%) resistant to ≥3 antibiotic classes. Notably, IV bag isolates tended to be more resistant than others (6/6 IV bag isolates compared to 1/8 others) (**Figure 2**). The identification of contaminated intravenous fluids represented a critical finding, as these fluids are directly administered to vulnerable neonates and provided a plausible mechanism for the high attack rates observed during the outbreak period.

Sampling of the hospital’s primary water sources (tanks and reservoirs) in November yielded no *K. pneumoniae* growth, confirming that contamination originated from ward-level infrastructure rather than the source water supply (26). Environmental sampling in December detected *K. pneumoniae* in labour ward sink surfaces, as well as neonatal ward sink surfaces, weighing scale, hand-washing soap, injection and intravenous fluids, indicating incomplete source control. Additional targeted interventions were implemented, including replacement of contaminated soap dispensers and enhanced cleaning of sink areas to address biofilm accumulation. In January 2024, environmental sampling detected *K. pneumoniae* in sink surfaces only, in both wards. Enhanced disinfection, drain decontamination and hand hygiene reinforcements were subsequently carried out. Final environmental sampling on March 10, 2024, yielded no *K. pneumoniae* growth from any sampled areas, and no new cases were identified after January 2024, marking the successful termination of the outbreak (26).

### Retrospective characterisation of the outbreak strains

To further understand the transmission dynamics and identify the causative agent behind the 2023 outbreak, we sequenced 148 isolates in total (**Table 1**, **Figure 3**): 66 clinical isolates from the outbreak period, 16 environmental samples collected between October and January 2023, 10 clinical isolates from January-February 2024, and 56 historical clinical isolates spanning 2012-2022 for contextualisation.

When we confirmed species identity at the genomic level, only 92 isolates (75%) were *K. pneumoniae*. The remainder comprised other members of the *Klebsiella* species complex and four *E. coli* (see **Table 1**). This discrepancy between culture-based and genomic identification proved particularly striking among the 66 (n=57 passed quality control (QC) thresholds) clinical isolates from 2023 that had been culture-confirmed as *K. pneumoniae*: only 39 (68%) were confirmed by whole genome sequencing to be true *K. pneumoniae*. Of 11 environmental isolates passing WGS quality thresholds, 8 were confirmed as *K. pneumoniae*. Following QC procedures that excluded two *K. pneumoniae* genomes failing quality thresholds, we obtained a final dataset of 90 high-quality *K. pneumoniae* genomes: 42 clinical isolates and eight environmental isolates from 2023-2024, and 40 historical isolates from 2012-2022 (**Table 1**, **Supplementary File 3**).

Whole genome sequencing clarified that the 2023 neonatal sepsis outbreak was driven predominantly by a single clonal lineage, sequence type ST39 (**Figure 4**, **Figure 5**). Among the 42 *K. pneumoniae* clinical isolates from 2023 to 2024, 29 (69%) belonged to ST39, first appearing in June 2023 and persisting through January 2024. During this 7-month period, ST39 accounted for 29 of 51 sequenced neonatal sepsis isolates (57%) and was not identified in isolates from other age groups.

Examining the genetic relationships among ST39 isolates through phylogenetic analysis of core genome SNPs, 28 of the 29 ST39 isolates formed a remarkably tight transmission cluster, differing by no more than five SNPs from one another (range: 0-5 SNPs), a pattern consistent with clonal spread within the hospital setting (**Figure 5**, **Supplementary Figure 2A&B**, **Supplementary File 4**). These genetically clustered isolates had been collected between June 9, 2023, and January 23, 2024, spanning 32.7 weeks (229 days), and comprised only a single non-neonatal isolate (isolate 38614B1).

The ST39 outbreak clone exhibited a remarkably uniform genomic profile across all 29 isolates in the transmission cluster (**Supplementary Figure 3**). Twenty-eight of the 29 isolates carried the KL62 capsular locus and all 29 isolates carried the OL2α.2 O locus, and harboured yersiniabactin (*ybt* 15) associated with the ICE*Kp11* integrative conjugative element. The clone lacked hypervirulence markers, with hypermucoidy markers *rmpA* and *rmpA2* absent from all isolates, consistent with the lack of *Klebsiella pneumoniae* virulence plasmid.

### Plasmid and chromosomal resistance architecture of the ST39 outbreak clone

Most outbreak isolates (26/29, 90%) carried *bla*_CTX-M-15_, explaining the observed cefotaxime resistance. All outbreak isolates carried additional AMR genes that could explain other observed resistances, including *aac*(*3*)*-IId* (gentamicin resistance), *catA1* (chloramphenicol resistance), *tet(A)* (tetracycline resistance), *dfrA14*+*sul1*+*sul2* (trimethoprim-sulfamethoxazole resistance) (**Figure 5**).

In 25/26 *bla*_CTX-M-15_-positive strains, *bla*_CTX-M-15_ and *aac*(*3*)*-IId* were located on a 187 kbp IncFIBK-IncFIIK plasmid (designated pNS39_A, GenBank accession PX620516), conferring resistance to cefotaxime and gentamicin which are the WHO-recommended drugs for treatment of neonatal sepsis (**Supplementary File 6**). BLAST analysis identified *K. pneumoniae* strain INF168 plasmid pKP91 (accession NZ_LR890472.1) as the closest reference. In addition to pNS39_A, the 25/26 *bla*_CTX-M-15_-positive strains also shared an IncFIA(HI1) IncFIIK (66,876 bp) plasmid (designated pNS39_B); however, this plasmid did not harbour any AMR genes.

### Chromosomal integration of *bla*_CTX-M-15_ in a plasmid-free ST39 isolate

One isolate (38277B1) lacked plasmids entirely yet carried *bla*_CTX-M-15_ and *aac*(3)-IId within an 11.2 kb chromosomal resistance island of mosaic architecture, assembled through sequential independent insertion events rather than mobilisation as a composite transposon (see **Appendix B**). The island is bounded at its 3′ end by ISKpn14, an IS1-family element specific to *K. pneumoniae*, which bears a perfect 9 bp target site duplication indicative of recent transposition. The upstream boundary is marked by a novel unclassified IS element with no matching direct repeats, suggesting a more ancient or incomplete insertion event.

Comparative gene cluster analysis with reference isolate 38833B1—which carries *bla*_CTX-M-15_ on a plasmid—revealed that the chromosomal locus occupied by this resistance island in 38277B1 originally contained an approximately 35 kb Tn3-family transposon carrying *catA1* (chloramphenicol resistance). Flanking conserved chromosomal sequences mapping 46.2 kb apart in 38833B1 but only 11.2 kb apart in 38277B1 confirm a deletion insertion event at this site. A near-identical ISKpn14 copy (96% amino acid identity) in the downstream chromosomal flank of 38833B1 provides direct molecular evidence that ISKpn14-mediated homologous recombination drove this replacement: the ∼35 kb Tn3 transposon carrying redundant chloramphenicol resistance was excised and replaced by the compact resistance island conferring resistance to extended-spectrum cephalosporins (*bla*_CTX-M-15_) and aminoglycosides (*aac*(3)-IId)—a net antimicrobial resistance upgrade at a single chromosomal locus.

### Environmental sources and clinical linkage of ST39

Among the environmental *K. pneumoniae* isolates we sequenced, three samples from intravenous fluids proved to belong to the outbreak ST39-KL62 cluster. All three came from IV fluid bags that were actively in use at the time of collection: two samples collected in October 2023 (IF52E, IF55E) and one in December 2023 (IF26E). Phylogenetic analysis revealed these environmental isolates differed by only 1-3 SNPs from contemporaneous clinical isolates within the ST39 outbreak cluster (**Figure 5, Supplementary Figure 3**, **Supplementary File 4**).

Importantly, two of the three environmental ST39 isolates shared identical AMR genes and plasmid profile with the clinical outbreak strains, including the critical *bla*_CTX-M-15_ and *aac*(3)-*IId* pNS39_A: IF55E carried an identical 187 kbp plasmid, IF52E carried a slightly smaller (136 kbp) deletion variant. The third environmental isolate (IF26E) lacked any plasmids and did not carry *bla*_CTX-M-15_ or *aac*(*3*)-IId, but harboured identical chromosomally-located AMR genes as the other ST39 isolates (**Supplementary File 5**).

As noted above, neonatal sepsis case numbers declined following implementation of the intervention bundle in late October 2023, and the WGS data show this was associated with a decline in the ST39 cluster isolates specifically (**Figure 4**). No more *K. pneumoniae* cases were detected during February-March 2024, and no *K. pneumoniae* isolate was identified from environmental sampling beyond December 2023. Two (2/2) isolates identified by conventional microbiology as *K. pneumoniae* from sink surfaces in January 2024 turned out to be, in fact, non- *K. pneumoniae* by genomic analysis.

### Co-circulating clusters and genomic characterisation of historical isolates

In addition to the dominant ST39 outbreak, phylogenetic analysis identified two smaller putative transmission clusters among the 2023-2024 clinical isolates, involving independent *K. pneumoniae* strains and restricted to neonates (**Supplementary Figure 2A** and **Supplementary File 7**). The first cluster consisted of two genetically related isolates (typed as ST25 and ST25-1LV (a single locus variant) by MLST, separated by 10 SNPs) that were collected nine days apart in April 2023. Both isolates shared the KL2 capsule locus and plasmid-borne chromosomal *bla*_CTX-M_ genes (one carried *bla*_CTX-M-15_ on a 30 kbp plasmid and the other, *bla*_CTX-M-216_ on a 15 kbp plasmid) (**Supplementary File 6**). The second cluster comprised two identical ST1552 isolates (0 SNPs) collected six days apart in July 2023 (**Supplementary Figure 2B**). These isolates carried loci encoding the K25 capsular serotype, O1αβ O-antigen, yersiniabactin lineage 9 mediated by ICE*Kp3*. Notably, both isolates also harboured an identical 210 kbp IncFIB_K_ plasmid which carried *bla*_CTX-15_, along with *aac*(3)-IIa, *ant*(3’’)-Ia, *bla*_TEM-1B_1_, *dfr*A15, *qnr*S1 *sul*1 and *tet(*A) (**Supplementary Files 6 & 7**).

Additionally, two distinct transmission clusters of multidrug-resistant *K. quasipneumoniae* subsp. *similipneumoniae* were identified among the non-*K. pneumoniae* sensu stricto isolates (**Supplementary Figure 2C & D** and **Supplementary File 8**). Cluster 1 (ST334) comprised three isolates circulating over 19 days (27 March–14 April 2023), all harbouring the KL178 capsule locus and producing CTX-M-15 (harboured on large, multi-resistance 124 -246 kb IncHI1B_1_pNDM-MAR plasmids) (**Supplementary File 9**), with *Omp*K36 porin alterations (p.Ala183fs). Cluster 2 (ST425) consisted of three isolates collected within a 3-day period (13–15 May 2023) and displayed KL61 capsule locus, whilst also harbouring CTX-M-15 ESBL production. These were similarly carried on multiresistance 179 kbp IncFIB_K_/IncHI1B plasmids (**Supplementary File 9**).

Both clusters demonstrated identical *Omp*K36 porin mutations and shared plasmid-borne resistance to fluoroquinolones (*qnr*S1) and aminoglycosides (*aac(6’)-Ib-cr*). The temporal separation and genetic differences (distinct ST and KL types) between the two clusters, combined with identical ESBL genotypes, suggest independent introduction events of CTX-M-15-producing *K. quasipneumoniae* at this healthcare facility, with potential for rapid nosocomial dissemination as evidenced by the clustering of isolates within short timeframes.

Analysis of the 40 historical *K. pneumoniae* isolates identified considerable genetic diversity encompassing 29 unique sequence types, however ST39 was not detected (**Table 2**). ST1788 was most prevalent (5/40), followed by ST502 (4/40), ST147, ST37, ST86, and ST592 (2/40 each), with the remaining STs appearing only once. KL2 (8/40) and KL15 (5/40) were the most common capsular loci. KL62 was detected in four isolates (10%), each in a different ST background. ESBL genes were identified in 33% of the isolates (14/40), predominantly *bla*_CTX-M-15_ (13 out of 14 found), and no carbapenemases were found.

### Regional and global context of the outbreak strain

To contextualise the ST39-KL62 outbreak strain, we first compared our WGS data with those from a previously reported 2016 outbreak of ST39-KL23 in the neonatal unit at the Sir Edward Francis Small Teaching Hospital in Banjul, a coastal site 268 km from Bansang (**Figure 1A**) (57). Our analysis revealed that the 2023 ST39-KL62 outbreak strains were separated from the previous outbreak isolates by >1,000 SNPs, indicating distinct sublineages rather than persistence of the same clone within The Gambia.

To further contextualise the outbreak strain, we compared it with a global collection of 684 *K. pneumoniae* ST39 isolates from 50 countries across six continents (**Figure 6**, **Supplementary File 10**), representing all publicly available ST39 isolates in Pathogenwatch, as of 2 October 2025. The collection (available at https://pathogen.watch/collection/81f6uz2riu07-updatedst39globalcollection6nov25) showed distinct clonal structure with three main lineages, based on the LINcode classification system, which maps ST39 to sublineage (SL)39 (43). The most common clonal group (CG) within SL39, accounting for n=582/780 (75%) isolates, was CG39. This was followed by CG10192 (166/780) and CG10195 (32/780). The Gambian outbreak strains belonged to CG10192, which was dominant among African isolates but found in minor proportions in Europe, Asia, and the Americas, suggesting African origin of this lineage (**Figure 6**). In contrast, the earlier ST39-KL23 outbreak isolates from The Gambia were assigned to CG39 in this nomenclature system (22% African, predominantly international). CG39 isolates were mostly KL2 (339/582, 58%) and KL23 (230/582, 40%), collectively accounting for 98% of CG39 isolates. CG10192-KL62 genomes displayed by high ESBL prevalence (117/131, 89.3%), with *bla*_CTX-M-15_ as the exclusive ESBL gene detected. *bla*_CTX-M-15_ prevalence in this clone was particularly high in Africa (96.1%) compared to Asia (58.3%) and Europe (60.0%).

Carbapenemase genes were rare amongst CG10192-KL62 genomes (11.5%), with *bla*_NDM-1_ found in 14 of 134 African CG10192-KL62, and *bla*_NDM-5_ in one out of 12 Asian genomes (**Figure 6**).

## Discussion

Retrospective whole-genome sequencing of the outbreak isolates identified the transmitted strain as KL62-ST39 carrying plasmid-borne *bla*_CTX-M-15_ and *aac*(*3*)-IId and multiple chromosomally-encoded AMR genes, which was responsible for 57% of neonatal infections over 7 months, causing 29 neonatal deaths with 57% case fatality. The high case fatality likely reflects both pathogen characteristics and clinical capacity limitations, including diagnostic delays and restricted antimicrobial options, emphasising the need for comprehensive clinical care improvements.

In one of 26 *bla*_CTX-M-15_ -positive isolates (38277B1), *bla*_CTX-M-15_ and aac(3)-IId were chromosomally rather than plasmid-encoded, located within an 11.2 kb mosaic resistance island. Target site duplication analysis supports stepwise in situ assembly rather than mobilisation as a pre-formed unit (58, 59): ISKpn14 at the 3′ boundary retained a perfect 9 bp TSD consistent with recent transposition, while the novel IS at the upstream boundary lacked matching direct repeats (1/9 bp identity). The mosaic architecture—with aac(3)-IId flanked by Tn2-derived resolvase and transposase-like sequences on either side—is consistent with sequential, independent insertions building the dual-resistance module rather than co-mobilisation of *bla*_CTX-M-15_ and aac(3)-IId as a single unit. While this chromosomal configuration was observed in only one isolate, chromosomally-encoded resistance is not amenable to plasmid curing and is stably vertically inherited, representing an alternative route for resistance fixation within this lineage.

The outbreak strain was also identified in samples of intravenous fluids, with 1-3 SNPs between environmental and clinical isolates, confirming the original conclusion that this was the probable primary transmission source and highlighting critical vulnerabilities in medication preparation practices. The detection of multiple smaller transmission clusters beyond the dominant ST39 outbreak clone also indicates systemic infection control deficiencies requiring comprehensive, sustained interventions rather than isolated corrective actions.

The attempt to replace repeatedly punctured 100 mL bags with single-use 10 mL sterile water— effective during the acute response was not sustainable once emergency support ended. Recurrent stock-outs, higher per-dose costs, minimum-order requirements, and procurement delays through national supply chains led staff to revert to multi-use IV bags, re-creating the conditions for fluid-borne contamination. This experience underscores that IPC improvements relying on consumables will fail without parallel systems solutions: ring-fenced financing and forecasting, framework agreements with the medical stores, minimum buffer stocks on the ward, and practical alternatives (e.g., prefilled unit-dose ampoules, on-site aseptic compounding or closed-system transfer devices where feasible). Routine audit with simple indicators (proportion of doses prepared with unit-dose water; days of stock on hand) and rapid escalation pathways are essential to maintain gains after an outbreak is declared over.

The successful outbreak termination through a multifaceted intervention bundle—including infrastructure remediation, supply chain improvements, enhanced cleaning protocols, and staff training—demonstrates that effective control is achievable in resource-limited settings, though the 4-month interval to source control highlights implementation challenges. Priority areas for intervention include: (1) strengthening infection prevention infrastructure, including water systems, sterilisation equipment, and hand hygiene supplies; (2) establishing standard operating procedures for safe medication preparation with appropriate supplies; and (3) implementing sustainable genomic surveillance platforms for early outbreak detection.

Global contextualisation of the outbreak strain with other ST39 genomes revealed predominance of three specific ST39 lineages, CG10192-KL62 (present outbreak strain), CG39-KL23 (previously reported outbreak strain in a different Gambian neonatal unit (57)) and CG39-KL2 in African populations. CG39-KL23 appears to be globally distributed, acquiring diverse ESBL and carbapenemase genes in different locations. In contrast, CG10192-KL62 and CG39-KL2 were almost exclusively associated with *bla*_CTX-M-15_ and were more common in Africa than elsewhere. Notably, ESBL ST39 outbreaks have also been reported in neonatal units in East Africa, consistent with continental expansion of these *K. pneumoniae* clones and highlighting their clinical importance.

With antibiotic resistance rendering empirical regimens increasingly ineffective and infrastructure limitations hindering infection control, maternal vaccination targeting prevalent African serotypes—particularly KL62 and KL2—represents a potential intervention. Notably, both KL2 and KL62 fall within the top five, and KL23 in the top 10, most prevalent K locus types associated with neonatal sepsis in Africa and South Asia (11). Additionally, 81% of *K. pneumoniae* clinical isolates from 2023, and 69% overall, carried one of the top 10 K loci. This supports the feasibility of vaccine-based prevention strategies targeting regionally relevant capsular serotypes. Genomic surveillance, as demonstrated here, provides essential data to inform vaccine design and establish baseline surveillance for impact evaluation.

This study has some limitations that warrant consideration. First, our environmental sampling was limited in scope, timing, and comprehensiveness. We may have missed critical contamination sources or transmission pathways that could have enhanced our ability to definitively trace the outbreak source. Secondly, while we identified contaminated intravenous fluids as a probable major transmission source based on genomic evidence and epidemiological linkage, we could not definitively establish the precise origin or directionality of contamination— whether fluids were contaminated at the source, during preparation, during storage, or during administration. Multiple potential transmission routes, including healthcare worker hands and other environmental reservoirs, may have contributed to the outbreak but were not exhaustively investigated.

Finally, our genomic investigation was conducted retrospectively after the outbreak concluded, potentially missing earlier transmission events and precluding use of the genomics-informed findings to support decision making during the outbreak. It has been demonstrated in other settings that real-time sequencing, including via rapid ONT-based sequencing as used here, can be deployed as part of outbreak investigations to inform IPC interventions and support outbreak resolution (60). This kind of sequencing can in principle be undertaken within the hospital laboratory using mobile ONT MinION devices, or at regional reference laboratories; in either scenario capacity needs to first be established for routine use, so that it is ready to be deployed during outbreaks (61–64) This is particularly challenging in low-resource settings, where even blood culture and susceptibility testing may not be routine (e.g., in this study, blood cultures were supported by a research study and outbreak isolates were sent to a supporting laboratory to undertake antimicrobial susceptibility testing) (65, 66). This underscores the broader need to build capacity for clinical microbiology, together with genomics, to reduce mortality from neonatal sepsis and other healthcare-associated infections, and from antimicrobial resistant infections more broadly (67).

### Conclusions

This investigation illustrates both the devastating impact of healthcare-associated *K. pneumoniae* infections in vulnerable neonatal populations and the actionable insights genomic epidemiology provides for outbreak response in resource-limited settings. Sustained investments in infection prevention infrastructure, laboratory capacity, and genomic surveillance—combined with accelerated vaccine development—are essential to reducing the unacceptable burden of neonatal sepsis mortality in sub-Saharan Africa and other high-burden regions.

## Data Availability

All data produced in the present work are contained in the manuscript.

https://pathogen.watch/collection/81f6uz2riu07-updatedst39globalcollection6nov25

## Competing interests

The authors declare have no competing interests.

## Funding

The PVS study was funded by GAVI alliance, the Bill & Melinda Gates Foundation and the UK Medical Research Council. Support for the sequencing and analysis of K. pneumoniae was provided by the Gates Foundation (INV066981 to GM, INV069410 and INV077266 to KEH). The study was designed, analysed and implemented by the authors. The funders had no role in study design, data collection and analysis, decision to publish, or preparation of the manuscript. The conclusions and opinions expressed in this work are those of the author(s) alone and shall not be attributed to the funders. Under the grant conditions of the Gates Foundation, a Creative Commons Attribution 4.0 License has already been assigned to the Author Accepted Manuscript version that might arise from this submission. Please note works submitted as a preprint have not undergone a peer review process.

## Author contributions

Conceptualisation – initial idea, hypothesis development and overall research design: G.A.M., A.K.S., K.E.H. Data collection – conducting experiments, collecting samples and gathering or analysing data: G.A.M., W.O.A., I.O., O.B., R.S., S.A.-T., M.J., A.N., E.F.-N. Methodology – development or design of the methodology; selecting and designing the research methods: G.A.M., A.K.S., K.E.H. Supervision – G.A.M., A.K.S., K.E.H. Software – E.E.O. Writing – original draft – writing the initial draft of the manuscript: E.F.-N. Writing – review and editing: E.F.-N., A.B., W.O.A., I.O., O.B., R.S., S.A.-T., M.J., A.N., E.E.O., K.E.H., A.K.S., G.A.M.

---

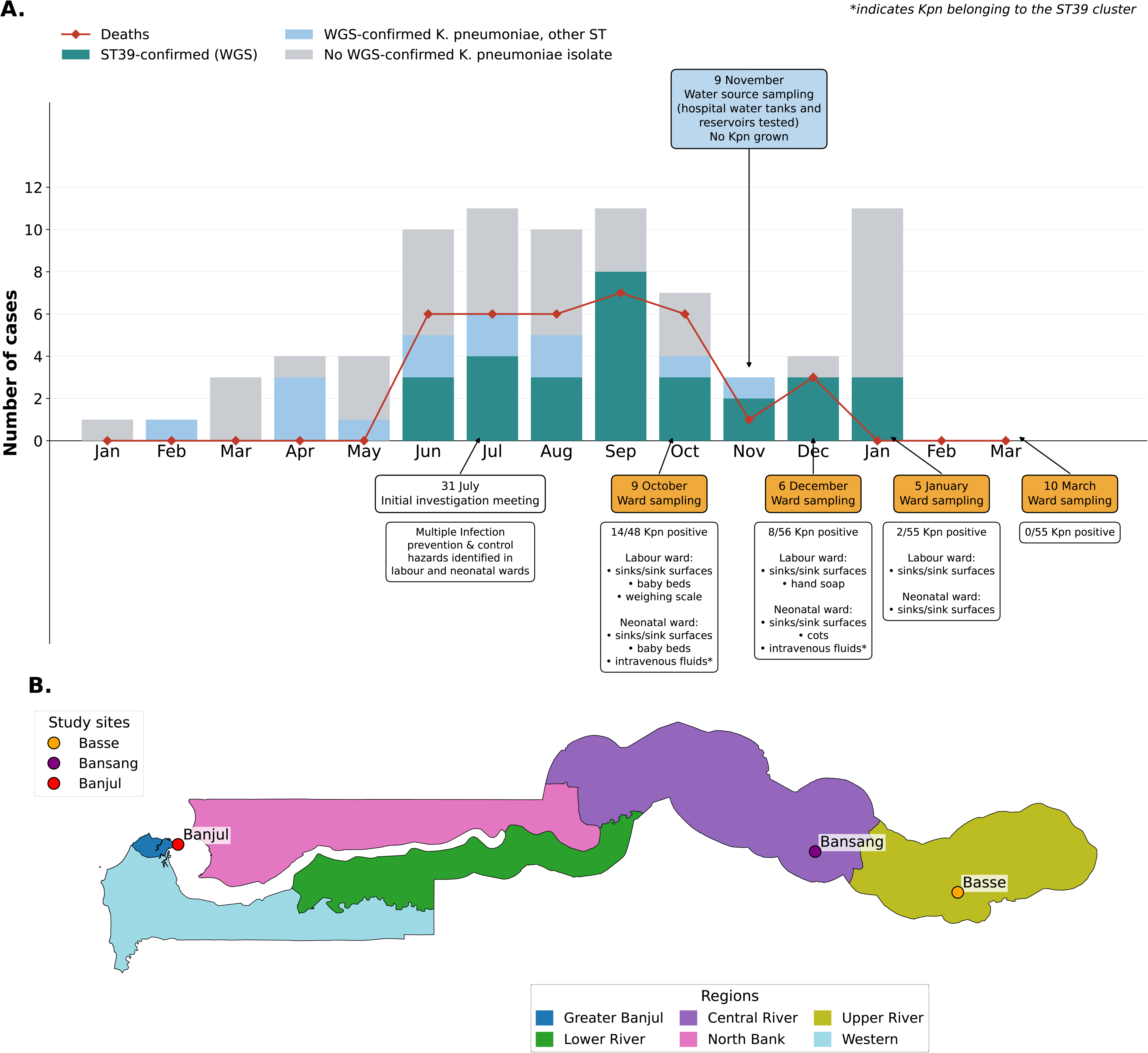

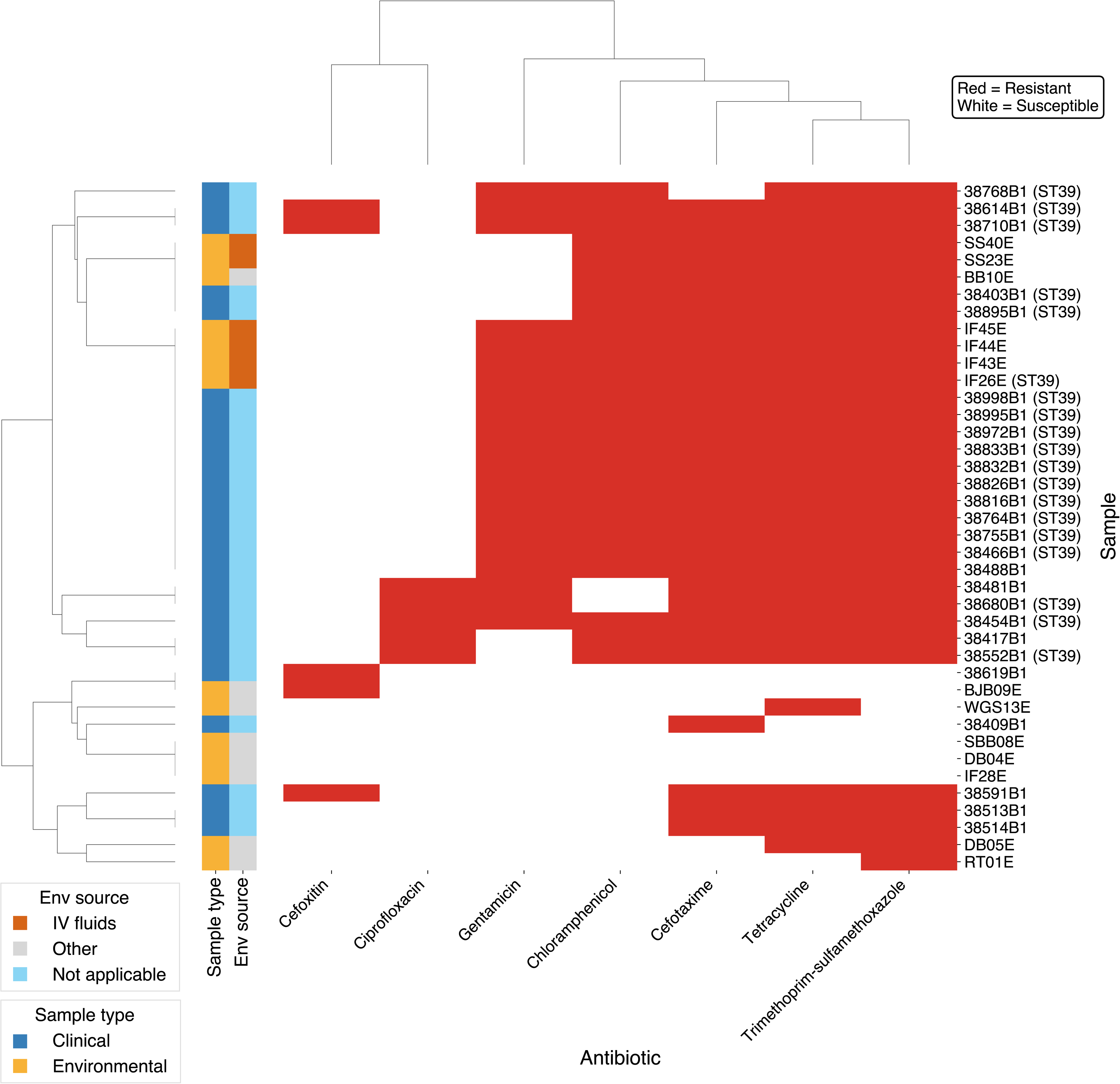

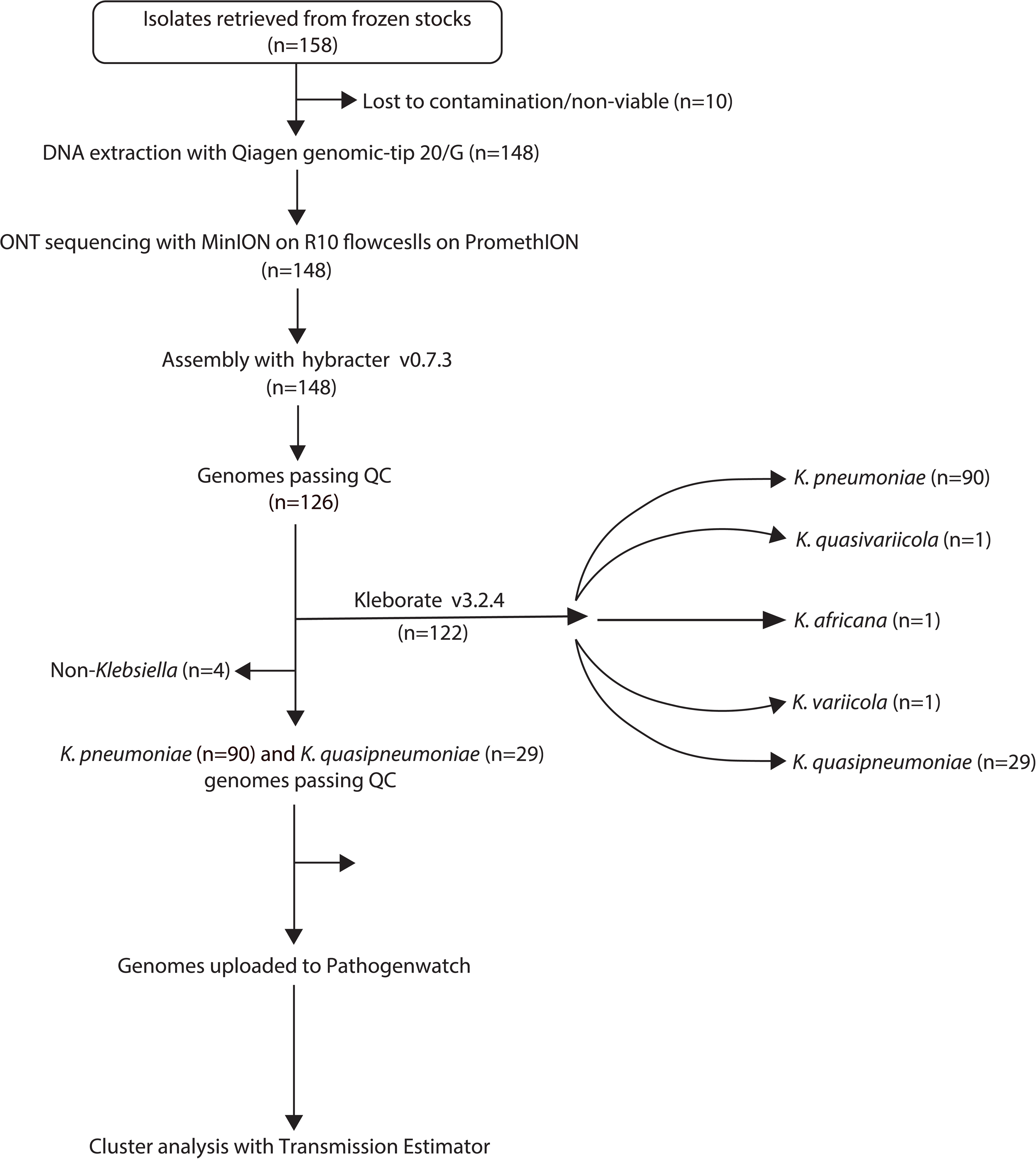

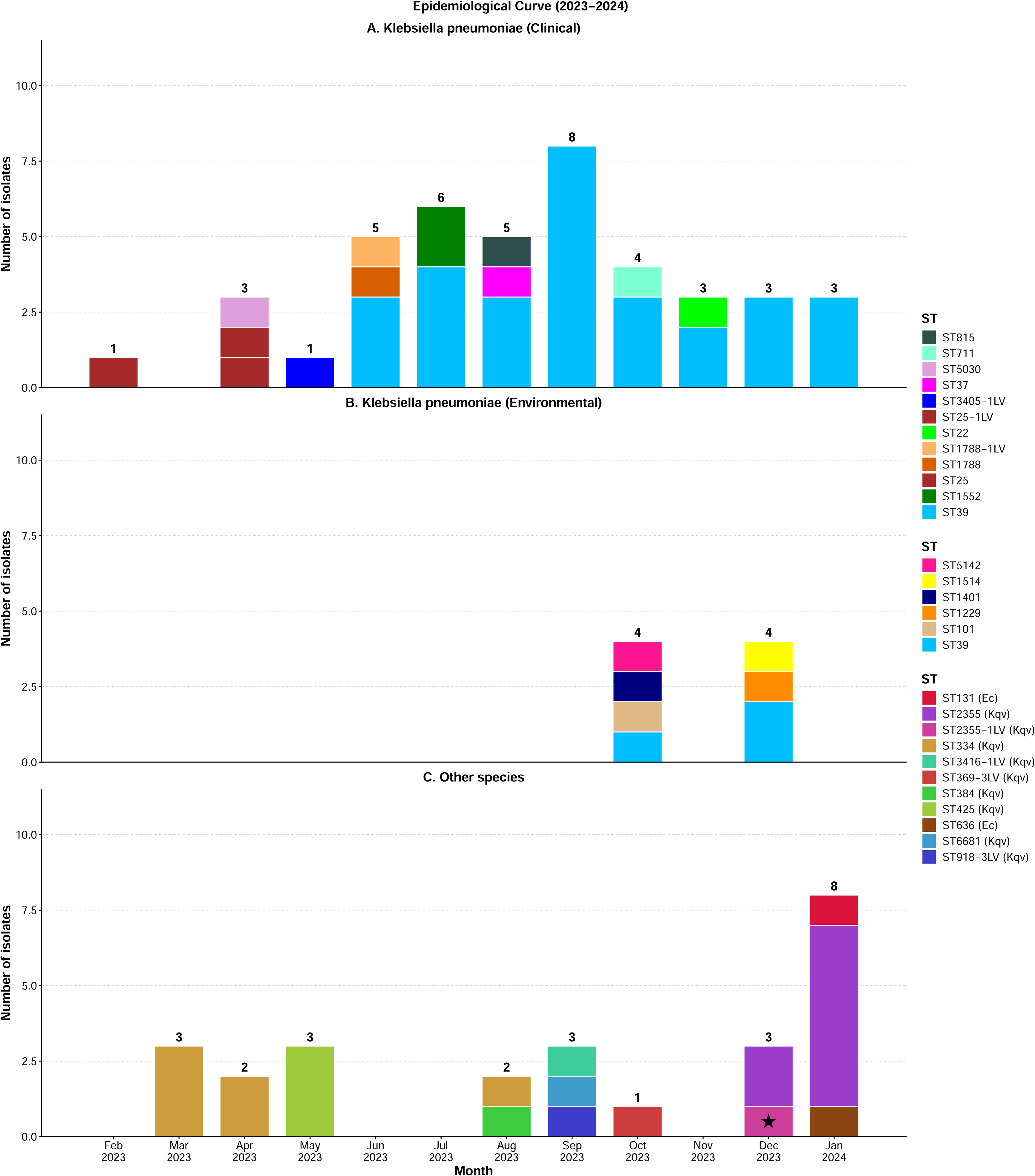

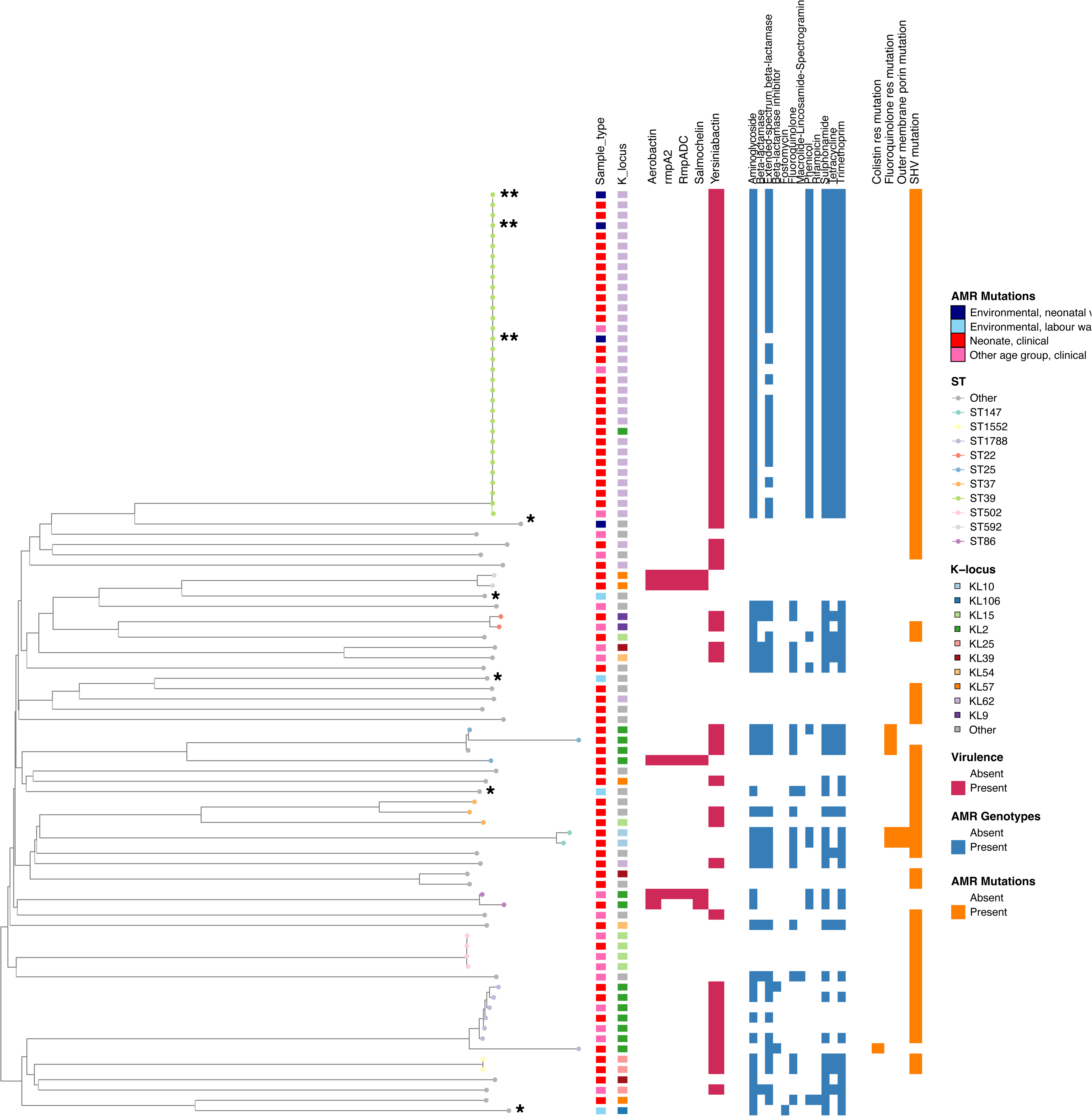

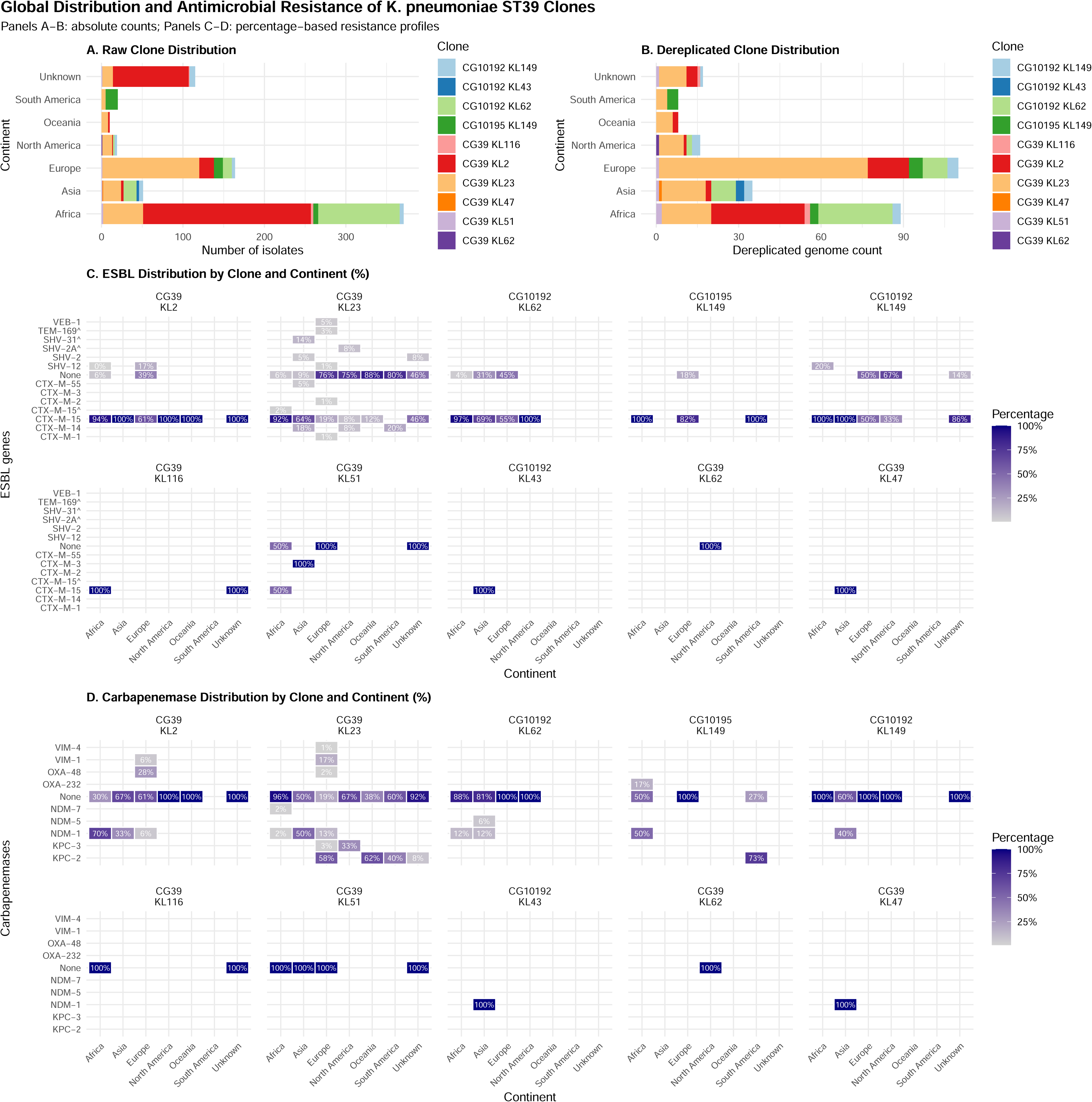

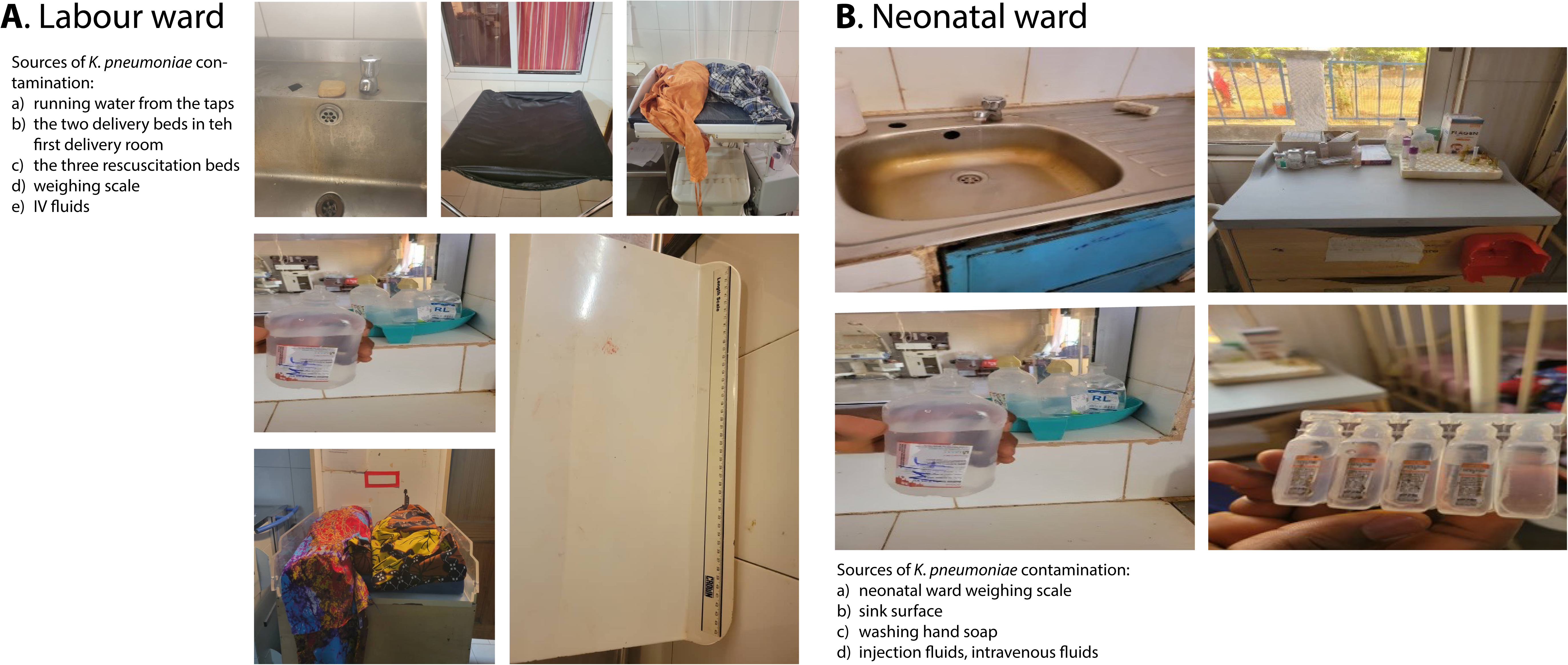

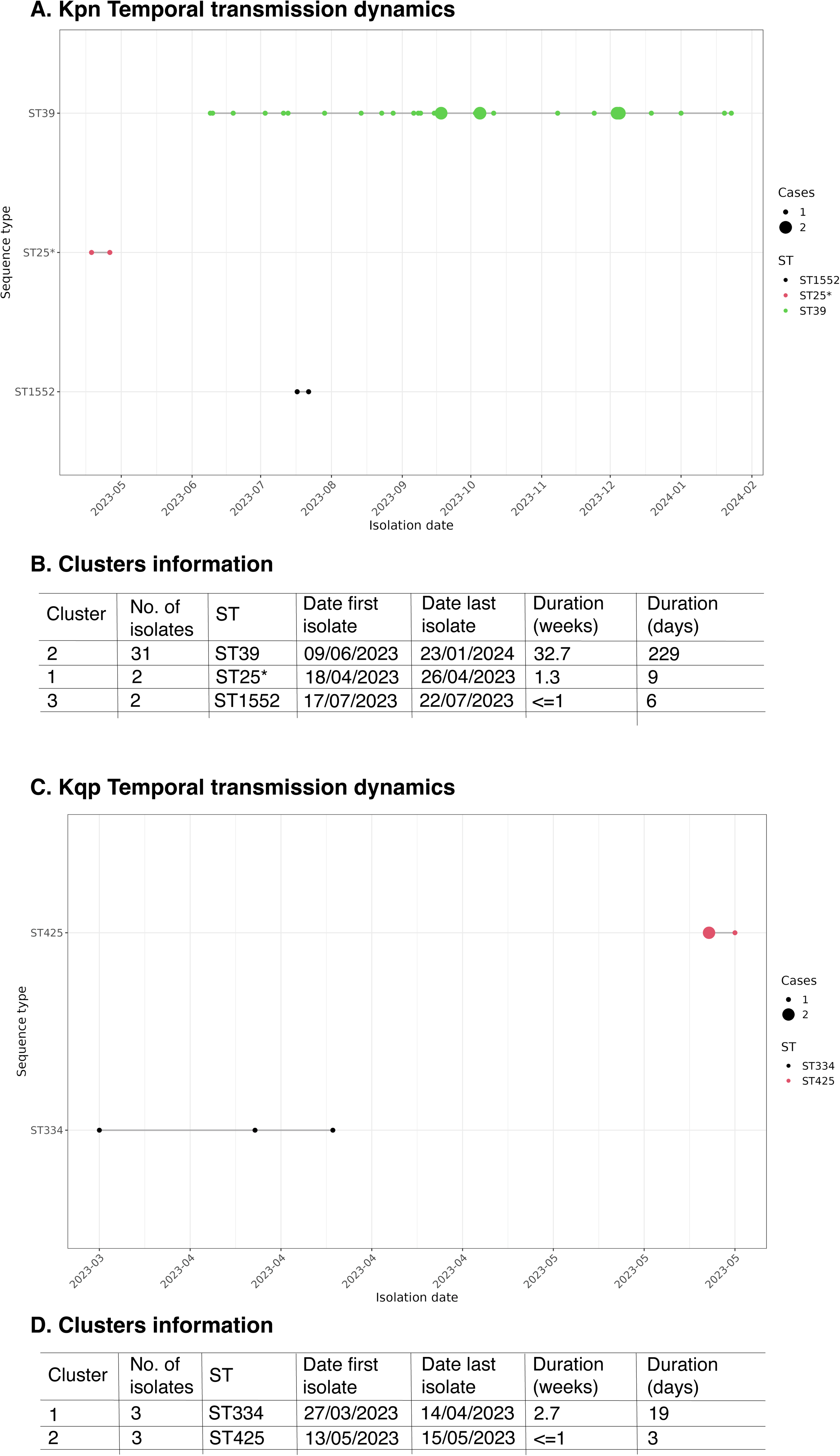

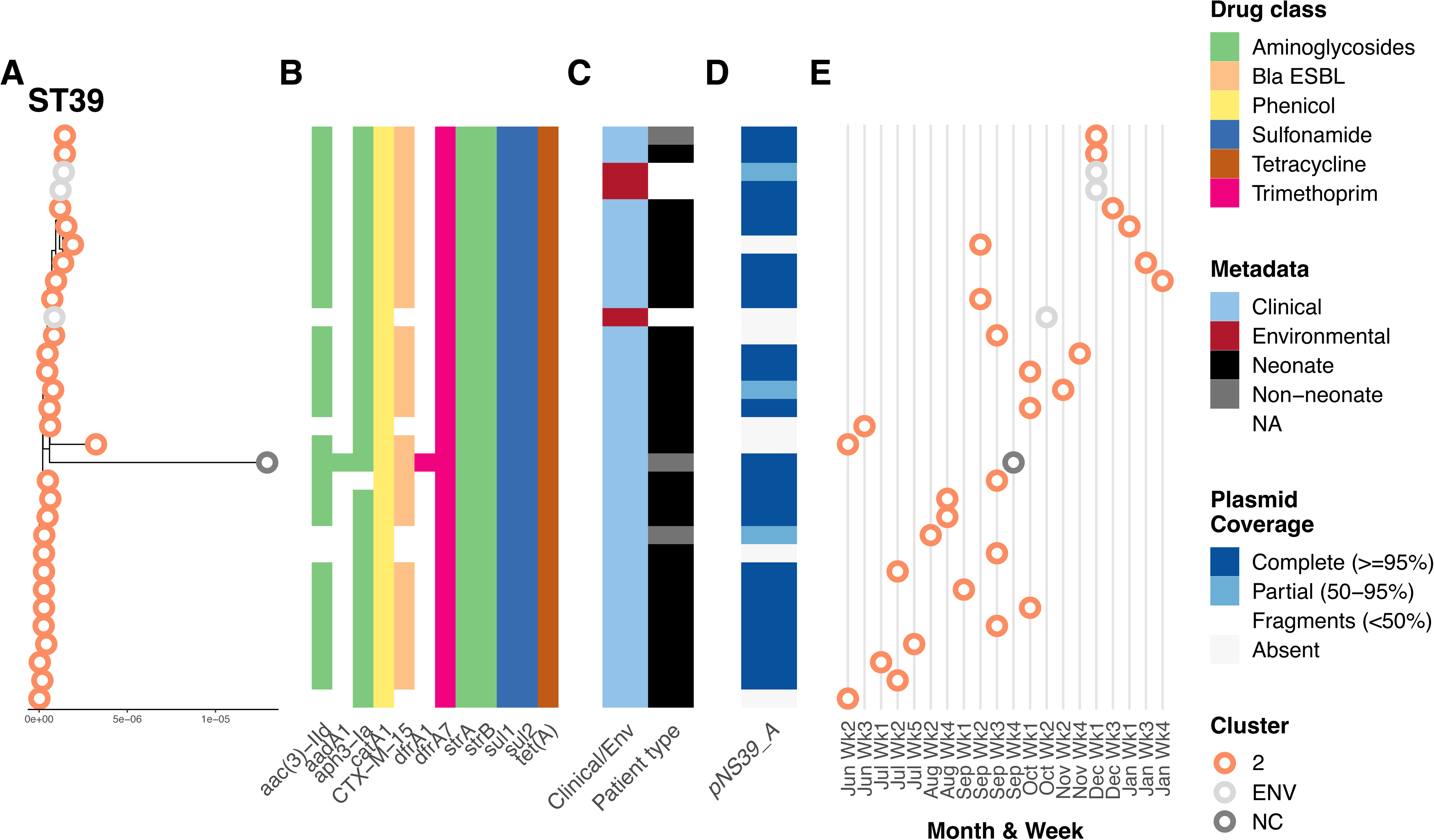

## Notes

### Competing Interest Statement

The authors have declared no competing interest.

### Author Declarations

The Gambia Government/MRC Joint Ethics Committee and the London School of Hygiene & Tropical Medicine Ethics Committee gave ethical approval for this work.

### Summary of Updates

The publication figures have been replaced with higher resolution ones.

