## Appendix A and Appendix B for "Molecular characterisation of a *Klebsiella pneumoniae* neonatal sepsis outbreak in a rural Gambian hospital: a retrospective genomic epidemiology investigation"

**A. Concordance beween Illumina and ONT sequencing data**

Recent studies have validated ONT sequencing for bacterial genomic surveillance and outbreak investigation. Evaluations of ONT R10.4.1 chemistry with *K. pneumoniae* demonstrate that assemblies with ≥35× coverage achieve <0.5% error rates for core genome multilocus sequence typing (1, 2), with deep learning variant callers enabling ONT to match or exceed Illumina accuracy even at 10× depth. Multiple studies confirm ONT-only data are suitable for high-resolution genotyping, serotyping, and phylogenetic clustering of K. pneumoniae using standard surveillance tools, with phylogenies stabilising after ~8 hours of sequencing (3, 4). While methylation-induced sequencing errors can affect cluster identification in some bacterial lineages, these can be addressed through bioinformatic masking or PCR-based library preparation (5).

To validate ONT-only data for our phylogenomic analysis, we benchmarked 17 samples sequenced on both platforms. Pairwise Pathogenwatch SNP distance analysis revealed high concordance between platforms**,** with identical samples differing by 0-14 SNPs (median: 4 SNPs, n=14/15 after excluding one outlier) (**Supplementary File 2).** Critically, identical assemblies generated on both platforms demonstrated strong topological concordance, clustering together with other isolates within the same outbreak cluster.

**B. Chromosomal integration of blaCTX-M‑15 in a plasmid‑free ST39 isolate**

One isolate (38277B1, 10 June 2023) lacked plasmids but carried *bla*_CTX-M-15_ and *aac(3)-IId* within an 11.2 kb chromosomal resistance region. BLASTn against ISfinder (6) identified the downstream IS element as IS*Kpn14* (768 bp, 100% identity, GenBank MH182641), an IS1-family element specific to *Klebsiella* *pneumoniae*, positioned 467 bp downstream of *bla*_CTX-M-15_. IS*Kpn14* created a perfect 9 bp target site duplication (TGAAGAGTC) at positions 95,125–95,133 and 95,902–95,910, consistent with IS1-family transposition mechanism (7), indicating recent IS*Kpn14* transposition at this locus.

Upstream of *bla*_CTX-M-15_, a 2.5 kb predicted open reading frame with homology to Tn3-family transposase (31% amino acid identity to TnpA_Tn3) separates *bla*_CTX-M-15_ from *aac(3)-IId* (639 bp and 338 bp intergenic gaps respectively). The arrangement—Tn2 resolvase (*tnpR*_Tn2, 542 bp) immediately upstream of *aac(3)-IId*, followed by a Tn3-family transposase-like ORF between *aac(3)-IId* and *bla*_CTX-M-15_—is consistent with *aac(3)-IId* having been inserted between the resolvase and transposase components of a Tn2-derived element, disrupting the ancestral Tn2 structure. The Tn2 remnant lacked *bla*TEM-1, which was identified on a separate contig (contig00049). A novel IS element (3,017 bp) on the opposite strand at the upstream boundary of the island could not be classified by ISfinder. Critically, the novel IS boundary lacked matching direct repeats (left: CACCACATT, right: TTACTGTCA; 1/9 bp identity), indicating the complete structure did not insert as a single unit (7).

The complete 11.2 kb structure comprises, in genomic order: (i) novel IS element (~3.0 kb, forward strand), (ii) Tn2 resolvase remnant (*tnpR*_Tn2, ~0.54 kb), (iii) *aac(3)-IId* (0.9 kb), (iv) Tn3-family transposase-like ORF (~2.5 kb), (v) *bla*_CTX-M-15_ (0.9 kb), and (vi) IS*Kpn14* (0.8 kb), all except the novel IS element on the complementary strand. This mosaic architecture indicates in situ assembly through sequential, independent insertion events rather than mobilisation as a composite transposon (6-8). IS*Kpn14* represents the most recent insertion event (preserved perfect TSD), while earlier insertions (novel IS, Tn2 remnant) have lost detectable TSDs or represent more ancient acquisitions (7).

**Mobile element replacement mediated by ISKpn14**

### To understand the evolutionary origin of this chromosomal integration site, we performed comparative gene cluster analysis with reference isolate 38833B1 (ST39-B, which carries *bla*_CTX-M-15_ on plasmid pNS39_A). Conserved flanking chromosomal sequences (92–100% nucleotide identity across six upstream and eight downstream genes) mapped 46.2 kb apart in 38833B1 but only 11.2 kb apart in 38277B1, confirming a deletion-insertion event at this locus (see **Figure** below).

BLAST analysis revealed that the replaced region in 38833B1 comprised approximately 35 kb of Tn3-related transposon sequences, including multiple Tn3-family transposases and TnAs3-like elements (91–100% identity to characterised transposons in Enterobacteriaceae databases) (9). This predecessor Tn3 transposon carried *catA1* (type A-1 chloramphenicol O-acetyltransferase, 99.85% identity, 100% coverage to NG_047582.1) (10), representing a duplicate copy in addition to the *catA1* present in the conserved ancestral chromosomal integron carried by all ST39 isolates.

Critically, comparative gene cluster analysis (see **Figure** below) revealed that an IS*Kpn14*-family transposase in the downstream conserved chromosomal flank of 38833B1 shares 96% amino acid identity with the IS*Kpn14* at the 3′ boundary of the 38277B1 resistance island. This near-identical IS*Kpn14* copy flanking the insertion site in the reference genome provides direct molecular evidence that IS*Kpn14*-mediated homologous recombination drove the transposon replacement event (8, 11): recombination between the resident IS*Kpn14* in 38833B1's chromosomal flank and the IS*Kpn14* associated with the incoming resistance island would have excised the ~35 kb Tn3 transposon and replaced it with the 11.2 kb mosaic resistance island in a single recombination event.

The replacement thus represents an antimicrobial resistance upgrade: a Tn3 composite transposon carrying outdated chloramphenicol resistance was supplanted by a compact 11.2 kb resistance island conferring resistance to frontline antibiotics (*bla*_CTX-M-15_: extended-spectrum cephalosporins (12); *aac(3)-IId*: aminoglycosides). This mobile genetic element exchange occurred without disruption of essential housekeeping genes, as both the replaced Tn3 transposon and the acquired resistance island are mobile genetic elements (8). The net deletion of 34,988 bp eliminated redundant chloramphenicol resistance (duplicate *catA1* copy lost; ancestral integron copy retained) while acquiring WHO therapy-defeating resistance.

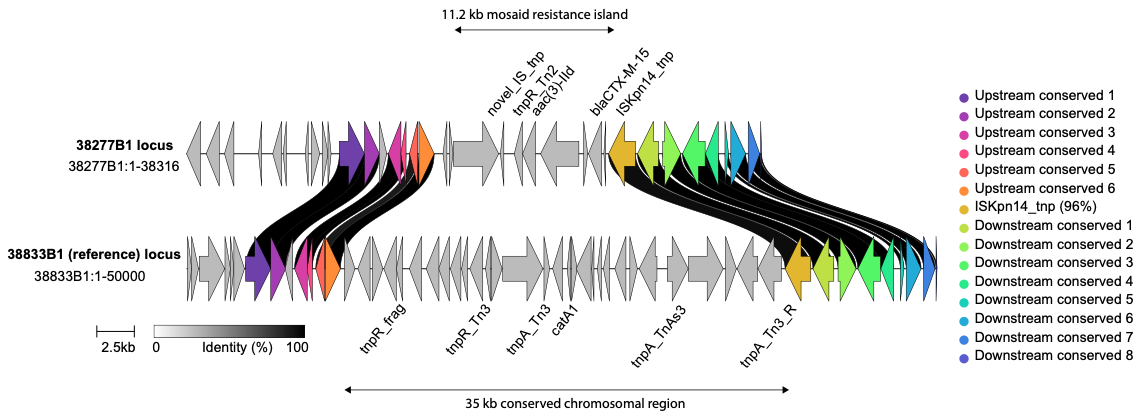

**Figure.** **Comparative** **genomic** **organisation** **of** **the** **chromosomal** **antimicrobial** **resistance** **locus** **in** ***K.*** ***pneumoniae* ST39** **strains** **38833B1** **and** **38277B1.** Gene cluster comparison generated with clinker (13)(Gilchrist & Tyson, 2021). Each horizontal track represents a chromosomal locus extracted from the reference strain 38833B1 (upper, complete genome) and outbreak strain 38277B1 (lower, draft assembly). Gene arrows are drawn to scale and coloured by sequence similarity group; arrows without labels represent conserved chromosomal flanking genes present at 100% nucleotide identity in both strains. Shaded ribbons between tracks indicate pairwise protein identity (colour intensity proportional to identity). Hypothetical proteins are unlabelled. In 38833B1, the locus is occupied by a ~35 kb Tn3 composite transposon carrying *catA1* (chloramphenicol acetyltransferase), bordered by multiple Tn3-family transposase (*tnpA*) and resolvase (*tnpR*) genes. In 38277B1, this transposon is replaced by an 11.2 kb mosaic resistance island comprising a novel insertion sequence transposase, a Tn2-family resolvase (*tnpR*_Tn2), aminoglycoside acetyltransferase *aac(3)-IId*, extended-spectrum β-lactamase *bla*_CTX-M-15_, and IS*Kpn14* transposase. An IS*Kpn14* copy (96% amino acid identity) present in the downstream chromosomal flank of 38833B1 is highlighted by a shared similarity ribbon, consistent with IS*Kpn14*-mediated recombination as the mechanism of transposon replacement.
